# Governance is key to controlling SARS-CoV-2’s vaccine resistance

**DOI:** 10.1101/2022.05.26.22275649

**Authors:** Kenichi W. Okamoto, Luis F. Chaves, Rodrick Wallace, Robert G. Wallace

## Abstract

Little attention has been paid to governance’s impacts on the evolution of SARS-CoV-2, the virus that causes COVID-19. To evaluate such impacts on the evolution of vaccine resistance, we analyzed a stochastic compartmental model to quantify the risk a mutant strain capable of evading immunity emerges post-vaccine rollout. We calibrated the model with publicly available data for four territories in the Western Hemisphere qualitatively differing in pandemic interventions. The model shows an immune-evading strain to be readily selected over all infectivities in Texas. In Panama, only a high level of transmission permits immune evasion to evolve. No invasion appears likely in Costa Rica and Uruguay. Programs combining pharmaceutical and nonpharmaceutical interventions are best positioned to remove the epidemiological space SARS-CoV-2 needs to evolve vaccine resistance.

**One Sentence Summary:** Modes of governance and production help set the evolutionary trajectories of vaccine resistance in SARS-CoV-2 before vaccine campaigns begin.

## Main Text

Considerable attention has been paid to public health’s impacts on the spread of SARS-CoV-2, the virus that causes COVID-19. Mask mandates, shelter-in-place orders, and broader interventions, including healthcare access and eviction moratoriums, mitigate SARS-2 transmission, albeit to varying degrees (*1*). Despite the striking emergence of multiple variants of concern, far less attention has been directed to policy’s effects upon the evolution of the virus (*2*). Particularly troubling is how little modeling has been dedicated to policy’s influence on the evolution of viral resistance to COVID-19 vaccines, for many policymakers our putative exit out of the pandemic (*3*).

Concerns about vaccine resistance may seem premature. Declines in vaccine effectiveness have indeed been documented across COVID variants, but the primary problem at hand still remains first getting millions around the world without vaccine access, atop those skeptical and hesitant, vaccinated (*2*). The issues of vaccine access and the evolution of resistance are intertwined, however. Delays, gaps, or reversals in vaccination and other interventions pursued in various combinations since the start of the pandemic have already set the trajectory for the evolution of immune evasion (*3*).

### Modeling governance’s impacts

To evaluate the possible impacts of governance on the evolution in vaccine resistance, our group analyzed a stochastic model to quantify the risk that a mutant SARS-CoV-2 strain capable of evading immunity emerges 170 days following vaccine rollout. The novel strain evades both natural immunity and immunity acquired by vaccination, but in this model otherwise circulates similarly as the ancestral strain. We calibrated the model with publicly available data for four territories in the Western Hemisphere: Panama, Costa Rica, Texas, and Uruguay. The four represent societies of relatively high Human Development Index (HDI), but also substantively different modes of governance and public health policy as they relate to responding to COVID-19.

We assessed how readily such a novel strain spreads in an initially monomorphic, immune-susceptible viral population. We found the ability of the strain to spread depends on the impacts of territorial policies on the ancestral strain’s epidemiology at the time the mutant emerges. While some of these effects are likely to be relatively constant, others are timevarying. The transmission coefficients of the ancestral strain on each day in each location are estimated using an Ensemble Kalman Filter (see Supplementary Methods and Figure S1) (*4*).

We then calculated the daily evolutionary invasibility of a mutant virus 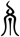 in each territory using a modified version of the basic reproductive number *R*_μ_ for the novel strain, derived using the next-generation operator (Supplementary Mathematica Notebook File S3) (*5*). We describe details of the modeling in the Supplementary Methods. All code is publicly available at github.com/kewok/immune_evasion and released under the GNU Public License (GPL 3).

Figure 1 shows SARS-CoV-2 exposure (and immunity) steadily low in Panama and Costa Rica across the 170 days after vaccinations begin, low but slowly rising in Uruguay, and sharply rising in Texas. The results also show that as the vaccination campaign begins, public health policies in Texas readily select for an immune-evading strain over all infectivities (found above the invasibility threshold log *R*_μ_ = 0). In Panama, only a high level of transmission 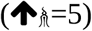 permits immune evasion to evolve. No invasion appears likely in Costa Rica. Even with rising exposure, resistance appears highly unlikely to successfully emerge in Uruguay.

**Fig 1.**
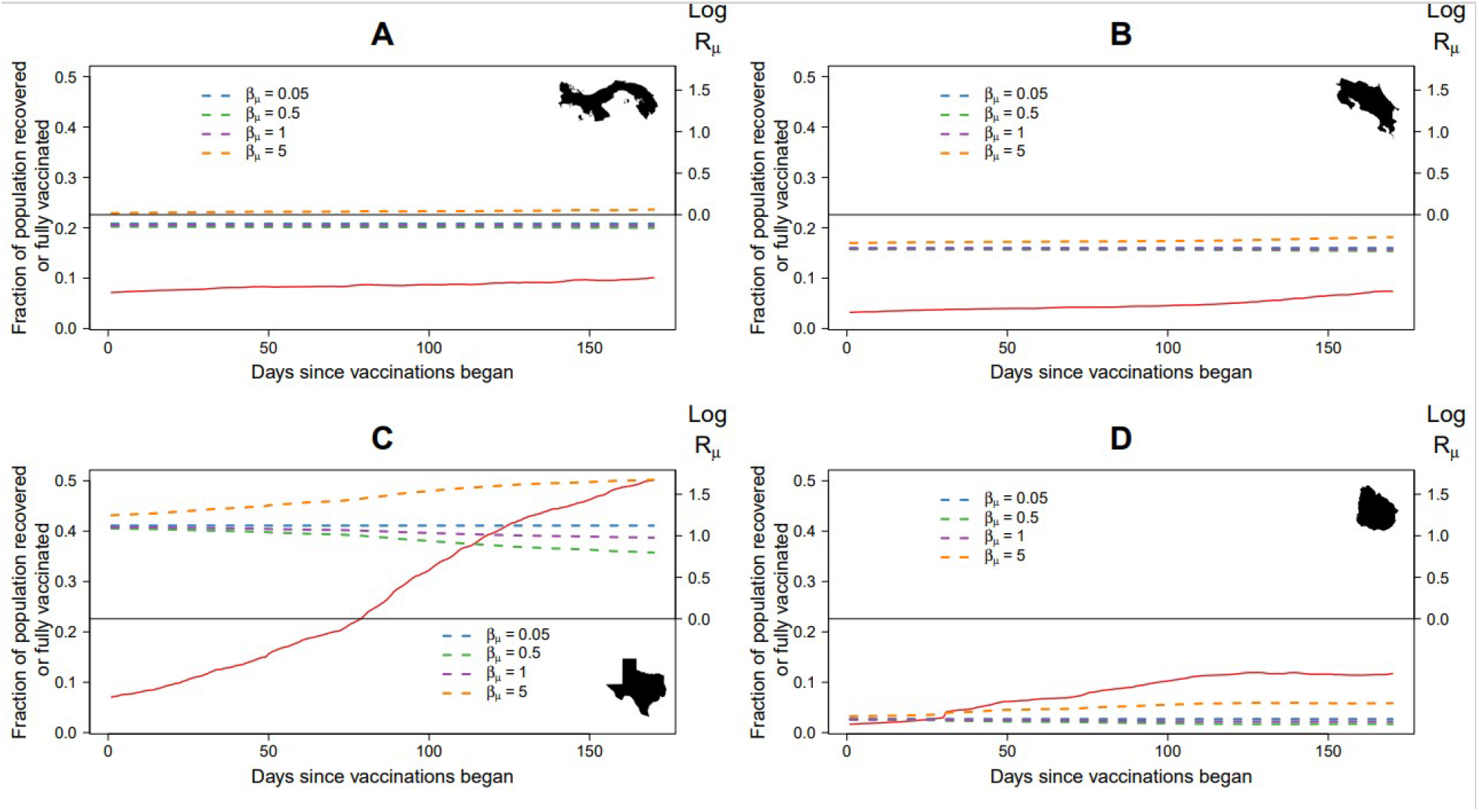
Exposure via infection or vaccination (solid line) and evolutionary invasibility (dashed lines) of an immune-evading mutant strain μ across 170 days in four Western Hemisphere territories with relatively high HDIs but qualitatively distinct public health strategies. (A) Panama, (B) Costa Rica, (C) Texas and (D) Uruguay. In all panels, the solid black lines represent the invasibility threshold (log Rμ = 0). The blue lines represent invasibility of a mutant 5% as infectious as the resident strain, the green lines the invasibility of a mutant half as infectious of the resident strain, the purple lines the invasibility of mutants as infectious as the resident strain, and the orange lines a mutant 500% as infectious as the resident strain.

The four territories differ substantively in their public health responses to the pandemic. The University of Oxford’s COVID-19 Government Response Tracker combines containment and closure policies, measures of population economic support, health system policies, and vaccine policies into a government response index (*6*). The index shows Panama, Costa Rica, and Uruguay engaged in the kinds of combinations of control efforts that U.S. COVID science leaders recently publicly denounced (*7*). To be sure, these campaigns are in flux. Panama eventually converged upon a full-spectrum response: masks, vaccines, and household support. Costa Rica’s nonpharmaceutical program pivoted to depending largely on vaccines alone beginning in 2021. Uruguay’s startling early successes were subsequently mitigated by demands to reopen the economy.

The Oxford effort also compiles subnational control programs, including for U.S. states (*8*). Texas’s responses were emblematic of an abandonment of COVID public health policies in the U.S. South, with minimal time under shelter-in-place, near-total reopening of businesses, and ending mask mandates despite new waves of COVID infection. Although Texas was among the first ten U.S. states to enact universal access to COVID vaccination (16 years and older), only 58% of Texans are fully vaccinated. That combination suggests SARS-CoV-2 populations there experience early and incomplete vaccine exposure and the granular geography of high- and low-vaccinated microhabitats to which resistance best evolves in response (*3*).

### Rethinking governance and science

Such arrays of pandemic-specific health policies appear to capture only a part of the variation in COVID population burden. One control theory model showed delays in responding alone degrade and destabilize governmental response capacity, whatever those interventions might be (*9*). The accumulating degradation promotes multiple infection waves of increasing severity, not unlike what we have seen across novel COVID variants. Dithering and denialism can amplify COVID’s damage beyond any immediate wave. The control system itself tailspins into long-term, unstable dynamics of repeated crashes and overcorrections. Under this class of models, the disease endemicity many hoped with the Omicron wave is replaced with explosive spikes of increasing amplitude.

The context of COVID-19 outcome can be expanded from public health responses to dominant modes of economic production. Across Mesoamerica and the Caribbean, clusters of countries for epidemiological variables such as the exponential growth rate, and COVID cases and deaths 100 days after detection of first case, were matched with clusters based on uneven development variables, including HDI, the WHO Universal Health Care index, and several trade indicators (*10*). In Mesoamerica and the Caribbean, trade openness was associated with increases in COVID cases and deaths. Increases in concentration of imports, a marker of uneven development, correlated significantly with increases in early epidemic growth and deaths, even while accounting for COVID mitigation and prevention polices, and human travel across countries.

The implications of these lines of analysis extend beyond governance and public health practice and into research itself. A disease outbreak, for instance, represents more than a convergence of susceptibles, the exposed and infected, and those who have recovered from infection. Public health interventions and traditional SEIR modeling from which such campaigns often take direction must reach beyond cataloguing the emergent properties of populations of interacting individuals (*11*). The broader social landscapes in which pathogens evolve drive disease outbreaks (*10, 11*). The causes for the evolution of diseases are found as much in the field of the social determinants of health as in the object of the pathogen or patient population.

We can take matters a step further. Modes of governance and production represent more than the ways a society impacts population health. They often embody a society’s dominant principles and the institutional cognition—how groups think—by which such values are turned into policies (*11, 12*). “Social determinants” of health in reality are *societal determinations* that are devised and acted upon by specific people (*13*). As our model suggests, statal and parastatal practices set many of the boundary conditions in time and space over which subsequent vaccine resistance in COVID-19 and any pandemics to follow likely emerge, long before any actual vaccination begins. More proactive and structural approaches to public health interventions must therefore be pursued to exit the rolling COVID trap and prevent novel pathogens from emerging out of the spillover queue.

Here are two strategic objectives to start. First, implement a program of integrated pharmaceutical and nonpharmaceutical interventions that removes the epidemiological space SARS-CoV-2 needs to evolve vaccine resistance and other adaptive life history changes (*14*). If not the Zero COVID programs some countries have instituted, governments and communities should implement timely, consistent, and collaborative programs across ministries or affinity groups that drive SARS-CoV-2 under its local rates of replacement.

Second, with the bigger picture in mind, governance must pivot back to supporting modes of production such as agroecology, community-controlled forestry, and open medicine that, on the front end of disease emergence, retard new pathogens from spilling over into human populations, and, on the back end, place public health and economic development on mutually supportive trajectories (*15*). Whatever the principles around which they organize themselves, societies across the globe are otherwise unlikely to flourish in the face of the epidemiological and climatic consequences of acting as if they are little more than a stock market with a country attached.

## Data Availability

All data produced are available online at github.com/kewok/immune_evasion

## Acknowledgments

The authors declare no financial or non-financial competing interests. The present work was conducted independently of any funding the authors have received.

## Author contributions

Each author’s contribution(s) to the paper are listed here under the CRediT model.

Conceptualization: KWO, LFC, RW, RGW

Methodology: KWO, LFC, RGW

Investigation: KWO, LFC, RGW

Visualization: KWO, LFC, RGW

Funding acquisition: N/A

Project administration: KWO, LFC, RGW

Supervision: KWO, LFC, RGW

Writing – original draft: KWO, LFC, RGW

Writing – review & editing: KWO, LFC, RW, RGW

## Data and materials availability

All data are available in the main text or the supplementary materials.

## Supplementary Materials

Supplementary materials, including data, figure code, coding libraries, supplementary methods, and GNU license are available at https://github.com/kewok/immune_evasion.

Supplementary Methods Fig. S1

