## Supplementary Figure 1 for "Governance is key to controlling SARS-CoV-2’s vaccine resistance"

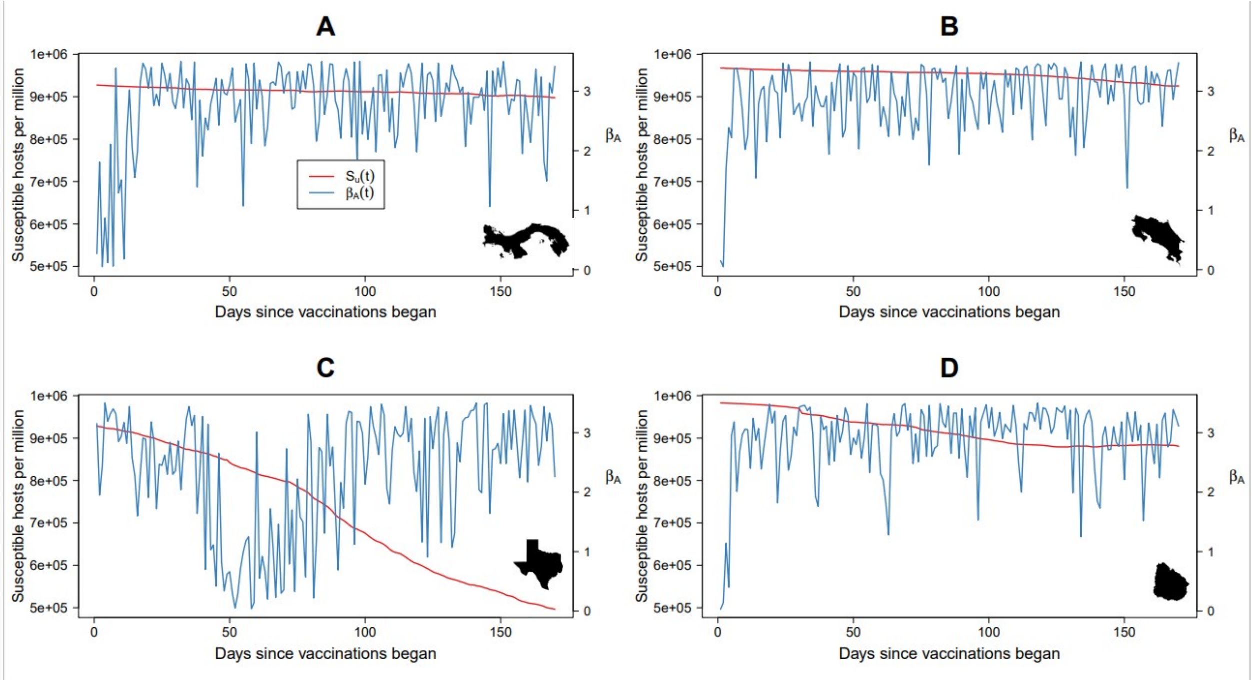


**Fig. S1.** Susceptible hosts per million (red lines) and the estimated daily infection coefficient $\beta_A(t)$ (blue lines) of the ancestral strain A estimated from the Ensemble Kalman Filter routine described in the Supplementary Methods for the four territories. The time horizon is as in Figure 1 of the main text: (A) Panama, (B) Costa Rica, (C) Texas and (D) Uruguay.
