## Supplementary Methods for "Governance is key to controlling SARS-CoV-2’s vaccine resistance"

Here we describe in further detail the modeling approach we use to assess the evolutionary invasibility of an immunity-evading strain $\mu$ of SARS-CoV-2 in a host population in which an ancestral, immune-susceptible strain $A$ circulates.

In the absence of vaccination or epidemiologically-relevant variability in the human or viral populations, the ancestral viral strain is assumed to circulate in the host population following an S$\rightarrow$E$\rightarrow$I$\rightarrow$R (Susceptible-Exposed-Infectious-Recovered) compartmental model over the time horizon (170 days following the roll out of vaccinations) that we investigate. The ancestral strain spreads through frequency-dependent transmission with a per-capita force of infection equal to $\beta_A I/N$, where $N$ is the host population ($N = S + I + E + R$) and $\beta_A$ the ancestral strain's infection coefficient. Based on Bi et al. (2020) and Qin et al. (2020), we use an incubation period $\sigma$ of 8.29 days and an infectious period $\gamma$ of 5 days. For purposes of the present analyses, we consider disease-induced mortality to not substantively alter the pathogen's dynamics, and that the total host population is approximately constant over the timescale of concern.

We then relax the assumption of immunity being acquired solely via infection by allowing susceptible hosts to become vaccinated. Thus, our S$\rightarrow$E$\rightarrow$I$\rightarrow$R model is modified so that vaccinated hosts are diverted into a compartment $V$, representing fully-vaccinated hosts. We assume fully-vaccinated hosts are unable to transmit the ancestral strain, but still circulate in the host population. Thus, after vaccinations begin, $N = S + I + E + R + V$.

Broadly, our basic strategy is as follows. First, we estimate the daily infection coefficient $\beta(t)$ for the ancestral strain $A$ in each polity from daily ($t$) data on incidence, prevalence and vaccination using an Ensemble Kalman Filtering regime. Next, we derive the average number $R_\mu$ of secondary infections by single, immune-evading mutant strain $\mu$ arising from a single host in which strain $A$ has mutated to strain $\mu$. $R_\mu$, in turn, is a time-dependent quantity whose magnitude depends, in part, on the ancestral strain's infection coefficient $\beta(t)$ at time $t$. Thus, we characterize how the invasibility of the mutant strain $\mu$ into the host population changes depending on (i) the phenotypic consequences of the mutation events giving rise to $\mu$, and (ii) the epidemiological profile (number of susceptible hosts, vaccinated hosts, etc...) of the host population when the mutant emerges. Below, we explain each of these in greater detail, as well as the `R` code used in our analyses. All underlying code is released under the GNU Public License v3 and is freely available at kewok.github.com.

#### Estimating the daily infection coefficient $\beta(t)$ from epidemiological data

Daily data recording total population size, as well as incidence, cumulative incidence, fatalities from SARS-CoV-2, as well as vaccinations against SARS-CoV-2 were collated for four polities in the Western Hemisphere (Panama, Costa Rica, Uruguay - collated as described in Chaves et al. 2020 - and Texas https://dshs.texas.gov/coronavirus/AdditionalData.aspx (https://dshs.texas.gov/coronavirus/AdditionalData.aspx); the underlying data are available at kewok.github.com). Thus, for a given polity **countryID**, the data are read in as a variable $d$

In [ ]:
```
d <- read.csv(paste('LatinAmerica_Data/',countryID,'.csv', sep=''))
```

General model parameters are specified as:

In [ ]:
```
btest = seq(0.01,3.5,by=0.01)
sigma <- 1/8.29
gamma <- 1/5
ief <- gamma/sigma
```

For polities representing countries (Panama, Costa Rica and Uruguay), initial conditions for infectious ($I_0$) and recovered ($R_0$) hosts were determined as follows:

In [ ]:
```
I0 <- sum(diff(d$total_cases[(min(which(!is.na(d[,'new_vaccinations_smoothed'])))-14):(min(which(!is.na(d[,'new_vaccinations_smoothed'])))) ])) # Prevalence based on all infectious from 14 days ago through present.
R0 <- d$total_cases[min(which(!is.na(d[,'new_vaccinations_smoothed'])))-14]
d <- d[min(which(!is.na(d[,'new_vaccinations_smoothed']))):nrow(d),]
d$Date <- as.Date(d[min(which(!is.na(d[,'new_vaccinations_smoothed']))):nrow(d),'date'],'%m/%d/%Y')
```

while for Texas, the initial conditions were specified as:

In [ ]:
```
I0 <- sum(diff(d$total_cases[(min(which(!is.na(d[,'new_vaccinations_smoothed'])))-14):(min(which(!is.na(d[,'new_vaccinations_smoothed'])))) ])) # Prevalence based on all infectious from 14 days ago through present.
R0 <- d$total_cases[min(which(!is.na(d[,'new_vaccinations_smoothed'])))-14]
d <- d[min(which(!is.na(d[,'new_vaccinations_smoothed']))):nrow(d),]
d$Date <- as.Date(d[min(which(!is.na(d[,'new_vaccinations_smoothed']))):nrow(d),'date'],'%m/%d/%Y')
```

In both cases, the dataset is concatenated to include only data following the beginning of vaccinations, and it is assumed that two weeks (approximately the incubation period plus the infectious period) must pass before new cases are considered no longer infectious.

For all locations, following the onset of vaccinations, the initial number of exposed individuals ($E_0$) is assumed to be a proportion $\pi$ of $I_0$ initially infectious individuals, with $\pi \sim N(\gamma/\sigma, 0.25)$ (see Engbert et al. 2021), and the initial number of vaccinated individuals $V_0$ is given by the number of people fully vaccinated in the dataset or, in the case of Texas, 0. The total population size $N$ is determined from XYZ. Thus, $S_0 = N - E_0 - I_0 - R_0 - V_0$.

Once initial conditions are determined, a time-lagged per-capita vaccination rate is calculated, whereby the daily fraction $v(t)$ of susceptible hosts vaccinated changes over time. Because of differences in the data reported, the routine for calculating the per-capita vaccinatino rate is somewhat distinct for Texas and the other polities. We assume that is takes 30 days following the initial administration of vaccination for individuals to enter the vaccinated compartment.

In [ ]:
```
if(countryID=='Tejas_revised')
    {
    vaxRate <-  c(diff(d$people_fully_vaccinated[1:31])/(N-d$total_cases[1:30] - d$Cumulative.Fatalities[1:30] - d$people_fully_vaccinated[1:30]), d$daily_vaccinations[31:nrow(d)]/(N-d$total_cases[31:nrow(d)] - d$Cumulative.Fatalities[31:nrow(d)] - d$people_fully_vaccinated[31:nrow(d)]))
    }
else
    {
    # impute fully vaccinated data if missing
    for (i in 31:nrow(d))
        {
        if (is.na(d$people_fully_vaccinated[i]))
            {
            d$people_fully_vaccinated[i] <- d$people_fully_vaccinated[i-1]
            }
        }
    vaxRate <-  c(rep(0,30), d[31:nrow(d),'new_vaccinations_smoothed']/(N-d$total_cases[31:nrow(d)]-d$total_deaths[31:nrow(d)]-d$people_fully_vaccinated[31:nrow(d)]))
    }
```

These data are then used to seed an Ensemble Kalman Filter (EnKF) algorithm to determine the daily infection coefficient $\beta$. Our approach loosely follows the strategy described in Engbert et al. 2021. Briefly, using the initial conditions and daily vaccination rates, we use the Gillespie algorithm (Gillespie 1977) to simulate 100 stochastic realizations of an S$\rightarrow$E$\rightarrow$I$\rightarrow$R model with a daily varying vaccination rates to identify distinct epidemiological trajectories up to a given date $T$ for a given infection coefficient $\beta$ ranging from 0.01 to 3.5 in increments of 1/100. An EnKF is then applied on the 100 trajectories to estimate the ensemble mean score for each value of $\beta$ on each date for each polity. The following segment of code illustrates the implementation of the EnKF:

In [ ]:
```
# Implement the EnKF:
rho <- 10
dt <- 1
Nruns <- 100

numb = length(btest)
E = matrix(data=rep(0,(Time-1)*numb),ncol=numb,nrow=Time-1)
best = rep(0,Time-1)
b = btest[j]
Z = matrix(data=rep(c(S0,E0,I0,R0),Nruns),ncol=4,nrow=Nruns,byrow=T)
Z[,2] = round(Z[,2]*(1+rnorm(Nruns,mean=0,sd=0.5)))
dt = d$dt[t]

# Simulate ensemble
sim = seir_custom(N=N,I0=Z[,3],E0=Z[,2],R0=Z[,4], V0=V0, b=b,a=a,g=
g,Time=dt,runs=Nruns, vaxRate=vaxRate)
Z[,] <- as.numeric(as.matrix(sim[,paste('Var',1:4,sep='')]))

# Ensemble Kalman filter
H = c(0,0,1,1)
P = cov(Z)
K = P%*%H/as.numeric(H%*%P%*%H+rho)

Zprime = Z
for ( r in 1:Nruns ) {
    Zprime[r,] = round(Z[r,]- 0.5*K*as.numeric(t(H)%*%Z[r,] + t(H)%
*%colMeans(Z) - 2*d$total_cases[t+1])) # Note they use cumulative ca
ses in xinfer.R
    Zprime[r,1] = N - sum(Zprime[r,2:4]) - sim[r,'Var5']    # enforc
e N = S+E+I+R+V
    }

E[t,j] = E[t,j] + 1/(2*(H%*%P%*%H+rho))*norm(H%*%colMeans(Z)-d$total
_cases[t+1])^2 + 0.5*log(H%*%P%*%H+rho)


Z = Zprime

E.df = data.frame(time=t, beta=b, E=E[t,j])
```

where the underlying stochastic realization of SEIR+V is simulated by the function **seir_custom**, in which the **ssar** package (Zepeda-Tello and Camacho-García-Formentí 2016) enables us to apply the stochastic Gillespie simulation when the vaccination rate parameter $v(t)$ is itself time-varying.

In [ ]:
```
install.packages("devtools")
devtools::install_github("INSP-RH/ssar")


# The state change/propensity matrix corresponding to
# A <- c("b*S0*I0/n","u(t)*S0","sigma*E0","g*I0")
# should look like:

# S -1 -1 0 0
# E +1 0 -1 0
# I 0 0  +1 -1
# R 0 0 0  +1
# V 0 +1 0 0

v <- matrix(c(
 -1, -1, 0, 0,
 +1, 0, -1, 0,
 0, 0,  +1, -1,
 0, +1, 0, +1,
 0, +1, 0, 0
    ),nrow=5, byrow=TRUE)

seir_custom <- function(N, I0, E0, R0, V0, b, sigma, g, Time, runs,
vaxRate, complete_trajectory = FALSE)
    {
    X <- matrix(c(S0=N-I0[1]-E0[1]-R0[1]-V0, E0[1], I0[1], R0[1], V
0), nrow=1)
    parameters <- c('b'=as.numeric(b), 'n'=as.numeric(N), 'sigma'=a
s.numeric(sigma), 'g'=as.numeric(g))

    u <- function(t) vaxRate[round(t+1)]
    # The time-dependent propensity function now becomes:

    pfun <- function(t, X, parameters)
        {
        cbind(parameters['b'] * X[,1] * X[,3]/parameters['n'], u(t)
* X[,1], parameters['sigma']*X[,2], parameters['gamma']*X[,3])
        }

    res <- ssa(X, pfun, v, parameters, tmin=0, tmax=Time, nsim=runs,
plot.sim=FALSE)
    if (complete_trajectory)
        return(res)
    else
        return(res[(nrow(res)-runs + 1):nrow(res),])
    }
```

All calculations described above were performed on the Mangi cluster at the Minnesota Supercomputing Institute at the University of Minnesota, Twin Cities.

We dentified the date- and polity-specific infection coefficient $\beta(t)$ by determining with the minimum ensemble mean score for that date and polity.

In [ ]:
```
locations <- c('Panama','CostaRica','Tejas_revised','Uruguay')

btest_index <- 1:length(seq(0.01,3.5,by=0.01))

Days <- 170
E_vals <- list(locations)


for (countryID in locations)
{
E_vals[[countryID]] <- matrix(nrow=Days, ncol=length(btest_index))

for (Date in 1:Days)
    {
    for (j in 1:length(btest_index))
        {
        inname = sprintf('LatinAmerica_Data/LKbeta/E_%s_%s_%s.dat',c
ountryID, j, Date)
        E_vals[[countryID]][Date,j] <- read.table(inname, header=
T)$E
        }
    }
}

beta.df <- data.frame(nrow=Days*length(locations),ncol=3)

current_row <- 1
bvals <- seq(0.01,3.5,by=0.01)
for (countryID in locations)
{
for (Date in 1:Days)
    {
    beta.df[current_row, 1] <- Date
    beta.df[current_row, 2] <- bvals[which(E_vals[[countryID]][Dat
e,]==min(E_vals[[countryID]][Date,]))[1]]
    beta.df[current_row, 3] <- countryID
    current_row <- current_row + 1
    }
}
```

### Derivation of the Average Number $R_\mu$ of Secondary Infections for the Immune-Evading Mutant Strain $\mu$

We assume that virions within a single host infected with the ancestral strain $A$ undergo a mutation (or series of mutations) within the single host that leads to the host becoming infected by virions of a novel strain $\mu$. Until this host infects other hosts, all other infectious hosts continue to be infected by the ancestral strain $A$.

We consider a situation where there are $S_u$ hosts that have never been infected by either strain, $S_v$ hosts that have either been vaccinated or previously infected by, and are now immune to, strain $A$. We further neglect coinfection. Under these conditions, the average number $R_\mu$ of secondary infections for the mutant strain $\mu$ characterizes the evolutionary invasibility of the novel strain. This quantity can, in turn, be determined from the next generation operator. In particular,

$$R_\mu(t) = \frac{S_v(t)(q+\sigma)b\beta_A(t)\sigma_v + S_u(t)\sigma\beta_A(t)(q+\sigma_v)}{\sqrt{n(q+\gamma)(q+\sigma)(q+\sigma_v)(S_v(t)(q+\sigma)b\beta_A(t)\sigma_v + S_u(t)\sigma\beta_A(t)(q+\sigma_v))}}$$

where $\gamma, \sigma, N, \beta_A$ are as above, $\sigma_v, b$ are the incubation period and relative infectivity, respectively, of the immune-evasive strain.

To contrast the different evolutionary effects of public health interventions, we model a situation where hosts infected by the novel strain $\mu$ are potentially removed from the host population by a rate $q$. We assume that this rate $q$ is reflective of the testing intensities across different polities. Thus we highlight how the evolutionary invasibility $R_\mu$ of strain $\mu$ depends not only on the mutant and ancestral strain's phenotypes, but also on the prevailing commitment to effective public health surveillance across the polities we examined. For detailed derivations, we refer the reader to the Supplementary Mathematica code.

Taken together, the average number $R_\mu$ of secondary infections is therefore calculated as:

In [ ]:
```
Rm <- function(b, B1, sigmaV, q, su, sv)
    {
    (B1*sigma*(q + sigmaV)*su + b*B1*(q + sigma)*sigmaV*sv)/sqrt(B1*
(gamma + q)*(q + sigma)*(q + sigmaV)*(n)*(sigma*(q + sigmaV)*su + b*
(q + sigma)*sigmaV*sv))
    }
```

### Characterizing the Evolutionary Invasibility of Different Immune-Evading Strains $\mu$ During the First Six Months of the Vaccination Campaign across Polities

We systematically explored how distinct immune-evading strains with different epidemiological phenotypes can emerge in each polity. In particular, we varied the mutant transmission coefficient $b$ relative to the ancestral transmission coefficient $\beta_A$ across three orders of magnitude (from 1/20$^{\text{th}}$ transmissibility to 5x transmissibility); varying the novel-strain specific incubation period $\sigma_v$ had very little discernable quantitative effect (results not shown), so we present results when the ancestral and novel strain have equivalent incubation periods.

On a given date $t$ in polity `countryID`, we therefore calculated the evolutionary invasibility $R_\mu$ of the immune-evading mutant virus as follows:

In [ ]:
```
bRange <- bRange <- c(0.05,0.5,1,5)) # e.g.; relative infectivities of the mutant strain

# Calculate the invasability value for each date across
get_Rms <- function(sigmaV, q, dats_by_day, countryID)
    {
    R_ms <- matrix(nrow=nrow(dats_by_day), ncol=length(bRange))
    for (i in 1:length(dats_by_day[,'Date']))
        {
        for (j in 1:length(bRange))
            {
            R_ms[i,j] <- Rm(bRange[j], B1[1], sigmaV, q, dats_by_day[i,'Su'], dats_by_day[i,'Sv'])
            }
        }
    return(R_ms)
    }
```

For each polity, starting with 30 days after which vaccinations began (assuming a two-dose schedule followed by a two-week waiting period - e.g., Polack et al. 2020 and Baden et al. 2021), we calculated the number $S_v(t)$ of hosts that are immune as the sum of the number of fully-vaccinated hosts and the total number of cases on a given day $t$. We further assumed that instantaneous co-infection by the two strains is negligible, and that new cases from infection by the ancestral strain $A$ are unable for infection by the novel strain $\mu$. Thus, the number $S_u(t)$ of susceptible hosts on day $t$ was therefore calculated as the difference between the total population size of the polity and $S_v$ as well as total deaths (Supplementary Figure S1).

Based on the daily $\beta_A(t)$ values estimated from the EnKF routine (Supplementary Figure S1), and assuming polity-specific testing rates (obtained Hasell et al. Sci Data 7, 345 (2020) and from Johns Hopkins University's testing dataset; https://coronavirus.jhu.edu/testing/states-comparison (https://coronavirus.jhu.edu/testing/states-comparison) Accessed Aug. 24 2021) are approximately constant in each polity, we then applied the function `get_Rms` to determine the daily evolutionary invasibilities of the novel strain illustrated in Fig. 1 of the main text. The code for the producing the data underlying the figures is reproduced below.

In [ ]:
```
sigma_v <- 1

locations <- c('Panama','CostaRica','Tejas_revised','Uruguay')
# Final daily values of beta_A
B1s <- read.table('LatinAmerica_Data/LKbeta/betaVals_revised.dat',he
ader=T)
Rms <- list()
susceptibles <- list()
vaxes <- list()
beta_ests <- list()

counter <- 1

for (countryID in locations)
{
d <- read.csv(paste('LatinAmerica_Data/',countryID,'.csv', sep=''))

# remove all records from before people were fully vaccinated:
if (countryID != 'Tejas_revised')
    {
    non_vax_days <- 1:min(which(!is.na(d[,'new_vaccinations_smoothed
'])))
    d <- d[-non_vax_days,]
    }

# Determine the time-lagged vaccination rate; assume it takes 30 day
s to become fully vaccinated
if(countryID=='Tejas_revised')
    {
    for (i in 1:nrow(d))
        {
        if (is.na(d$people_fully_vaccinated[i]))
            {
            d$people_fully_vaccinated[i] <-  d$people_fully_vaccinat
ed[i-1]
            }
        # As there is a date in Texas with NA deaths
        if (is.na(d$total_deaths[i]))
            {
            d$total_deaths[i] <-  d$total_deaths[i-1]
            }
        }
    Sv <- d$people_fully_vaccinated + d$total_cases
    }
else
    {
    # impute fully vaccinated data if missing; note the non-Texas da
ta begin when vaccinations began.
    for (i in 31:nrow(d))
        {
        if (is.na(d$people_fully_vaccinated[i]))
            {
            d$people_fully_vaccinated[i] <- d$people_fully_vaccinate
d[i-1]
            }
```

```
        }
    Sv <-  c(rep(0,30), d[31:nrow(d),'new_vaccinations_smoothed']) +
d$total_cases
    }

Su <- d$population - Sv - d$total_deaths
B1 <- B1s[which(B1s[,'LK_ID']==countryID),'b_est']
beta_ests[[counter]] <- B1
dats_by_day <- data.frame(Date=1:Days, Su=Su[1:Days], Sv=Sv[1:Days])
n <- unique(d$population[1])
my_q <- tests_by_country[countryID] / (n/1e6) # Convert tests per mi
llion into tests per capita
susceptibles[[counter]] <- Su[1:Days] / n * 1e6 # Convert susceptibl
es to per-million
vaxes[[counter]] <- Sv[1:Days]/n # Convert vaccinated to per-million

Rms[[counter]] <- get_Rms(sigma_v, my_q, dats_by_day, countryID)
counter <- counter + 1
}
```
